# Anthropogenic Debris in Bivalves Sold for Human Consumption in Northern California: A Cross-Sectional Follow-Up Study

**DOI:** 10.1101/2025.07.03.25330794

**Authors:** Zaeda Esperanza Metcalfe, Jason Limberis, John Metcalfe

## Abstract

Microplastics have emerged as a major environmental concern due to their widespread presence and potential health impacts. Growing water contamination has raised concerns about its presence in seafood consumed by humans. In this single-center, cross-sectional study, we collected a sample of 10 Pacific oysters sold locally in Northern California. We compared microplastic content with a prior study performed at the same location in 2015 (Rochman et al.). Anthropogenic debris was extracted from the whole tissue of shellfish using a 10% KOH solution and quantified under a dissecting microscope. Microplastics were found in all ten samples (compared with 4/12 samples in 2015), from three to greater than 40 particles per sample. These anthropogenic debris were primarily filaments from 0.2 to 2 mm long. Our study findings suggest that microplastics in bivalves sold for human consumption have increased in the past ten years, with potential human health concerns.

## Background

Microplastics, plastic particles smaller than 5 mm, have recently emerged as a major environmental concern due to their widespread presence and potential health impacts. These tiny particles are now found worldwide, even in pristine environments, with oceans being among the most affected ecosystems. The leading causes of plastic debris in marine environments are urban development and population density, as runoff and waste discharge funnel plastics into waterways that lead to the ocean. This growing water contamination has raised questions about its prevalence in marine life, especially seafood consumed by humans.

Understanding the potential health risks microplastics pose to human health is a critical research priority. As global plastic production continues to rise, so does the urgent need to investigate the long-term effects of plastics pollution on ecosystems and human health.

We aimed to track changes in microplastic contamination in bivalves sold for human consumption locally in Northern California. Specifically, we compared the prevalence of microplastic debris in Pacific oysters to findings from a 2015 study^1^ that analyzed microplastics among fish and bivalves sold in Half Moon Bay.^1^ By following the same methods as the earlier study, we identify trends in microplastic accumulation and data to guide future efforts. Because microplastic amounts rise with population growth and urban development, we hypothesized that the prevalence of microplastics would have risen since 2015 when the previous experiments were performed.

## Methods

Ten Pacific oysters (*Crassostrea gigas*) were purchased from a fish market at Princeton Harbor, Half Moon Bay, California, and transported in a cooler. We replicated the analytical methods and sample processing of the Rochman 2015^1^ study, with the exception of particle measurement. In our study, we estimated anthropogenic debris size based on dissection microscope magnification rather than using a micrometer. Briefly, oyster samples were immediately processed for analysis. First, shells were removed and then the whole tissue of each sample was placed into individual jars. Wearing protective gear (gloves, goggles, and a lab coat), a 10% potassium hydroxide (KOH) solution was prepared, using a scale for precision, and added to each jar at three times the volume of the tissue. The jars were sealed and incubated at 60°C for 36 hours in a water bath inside an oven (simulating a bain-marie) to dissolve all organic tissue (but not plastics). To avoid contamination, negative controls (glass petri dishes containing only ultrapure water) were placed in the laminar flow hood during the extraction procedure and next to the microscope during specimen examination. A positive control of microplastics obtained from drill shavings of a rock-climbing hold subjected to KOH digestion was also analyzed. All tools and glassware were rinsed three times with ultrapure water before use. After digestion, samples were poured through a 0.2 mm sieve in a laminar flow hood (at a UCSF lab space) to prevent airborne contamination. The leftover material on the sieve was transferred to labeled glass petri dishes. A negative control petri dish containing ultrapure water was placed in the laminar flow hood to assess for contamination. Samples were examined using a dissection microscope (10x–40x). Petri dish covers were removed, when necessary, with a second negative control in the viewing area to assess for airborne contamination. Anthropogenic debris in each sample was identified by color, shape, texture, and uniformity, and a TRAPDOOR 3-Axis Digiscoping Phone Adapter with a smartphone was used to photograph findings.

## Results

Microplastics were found in all ten specimens (**Table**), with some samples containing more than 30 visible microplastics. The microplastics found were mostly filaments, with few fragments and films. We estimate that microplastics were 0.2-2 mm in length based on magnification and comparison with prior findings^1^. In negative control #1 (standing ultrapure water on our viewing station), there was one red filament hypothesized to be from a similar color napkin in the room. A similar red filament was found in oyster #8 and was considered contamination and excluded. In negative control #2, placed inside the laminar flow hood where digested material was being sieved and placed in petri dishes, we found a single clump of black filaments (**Figure S1**). These were hypothesized to be from the black gloves worn during the experiments. No similar debris was found in any of the samples. As a positive control, we utilized a packaged oyster purchased from Trader Joes, adding plastic toothbrush filaments and green climbing hold shavings. It was digested, like the others, to ensure that the digestion process only decomposed organic tissue and not the plastics; microplastics were readily observed after the experiment, as expected.

## Discussion

We show in a carefully controlled experiment that since 2015, the amount of microplastic contamination in Pacific oysters sold as food in Half Moon Bay, California, has increased. In the 2015 experiment, only four out of twelve Pacific oyster specimens had microplastics in them. Further, in those specimens, only one or two pieces of anthropogenic debris were found in each specimen. In contrast, 100% of our samples had microplastics, many with high amounts. Filaments, rather than fragments, films, or foam balls^2^, were by far the most common type of microplastic in our experiment, as in the 2015 study. As explained then, this could be because large concentrations of filaments end up in California’s aquatic habitats from wastewater outflows; although our wastewater management systems are advanced, synthetic fibers from washing machines can remain in sewage effluent. The 2015 study only considered particles greater than 0.5 mm. We expanded on this by examining all anthropogenic debris down to 0.2 mm, which could in part explain the increased levels of microplastics found in our study.

Nowadays, microplastics are everywhere. In fact, it is estimated that humans consume a “credit card” worth of microplastics each week.^2^ As the human population continues to grow, cities do as well, leading to microplastics entering our natural environments. Ocean currents, wind, or animals then spread anthropogenic debris. These tiny pieces of plastic are so small that they are airborne and can even enter our bodies through damaged skin^3^.

Since microplastics are ubiquitous, our experiment had to be very carefully controlled^4^. For example, we had to use ultrapure water instead of tap water, all instruments and containers had to be washed in ultrapure water multiple times, the air around the sample had to be clean which is why the extraction was done in the laminar flow hood, and we used multiple negative controls to capture any possible contaminant from the experimental environment. This field of study only becomes harder as more and more plastics are introduced to our natural environments.

The bivalves studied in this experiment were sold for human consumption yet contained substantial anthropogenic debris inside. This discovery raises a serious question: Are Pacific oysters and other bivalves still okay to eat? Also, if we found this debris in seafood, what about our other foods? Unfortunately, there has not been much research about this or in this field because of how difficult and tedious the processes are to obtain accurate results. Little is known about the health effects of microplastics on humans or other animals, though some studies suggest harm.^3^ Now that we know how abundant they are, the next step is to figure out their human health impacts in order to advocate for their removal and replacement with biodegradable plastics. That will be my goal.

My experiment had some limitations. I looked at only one species from one fish market at one point in time. For some samples, the digestion was not complete despite the prolonged duration; in these samples, my microplastic counts were likely an underestimate. Finally, we examined microplastics down to 0.2 mm; smaller microplastics (≤0.01 mm) are likely more threatening to human health, but difficult to detect with current methods.

**Table 1.**
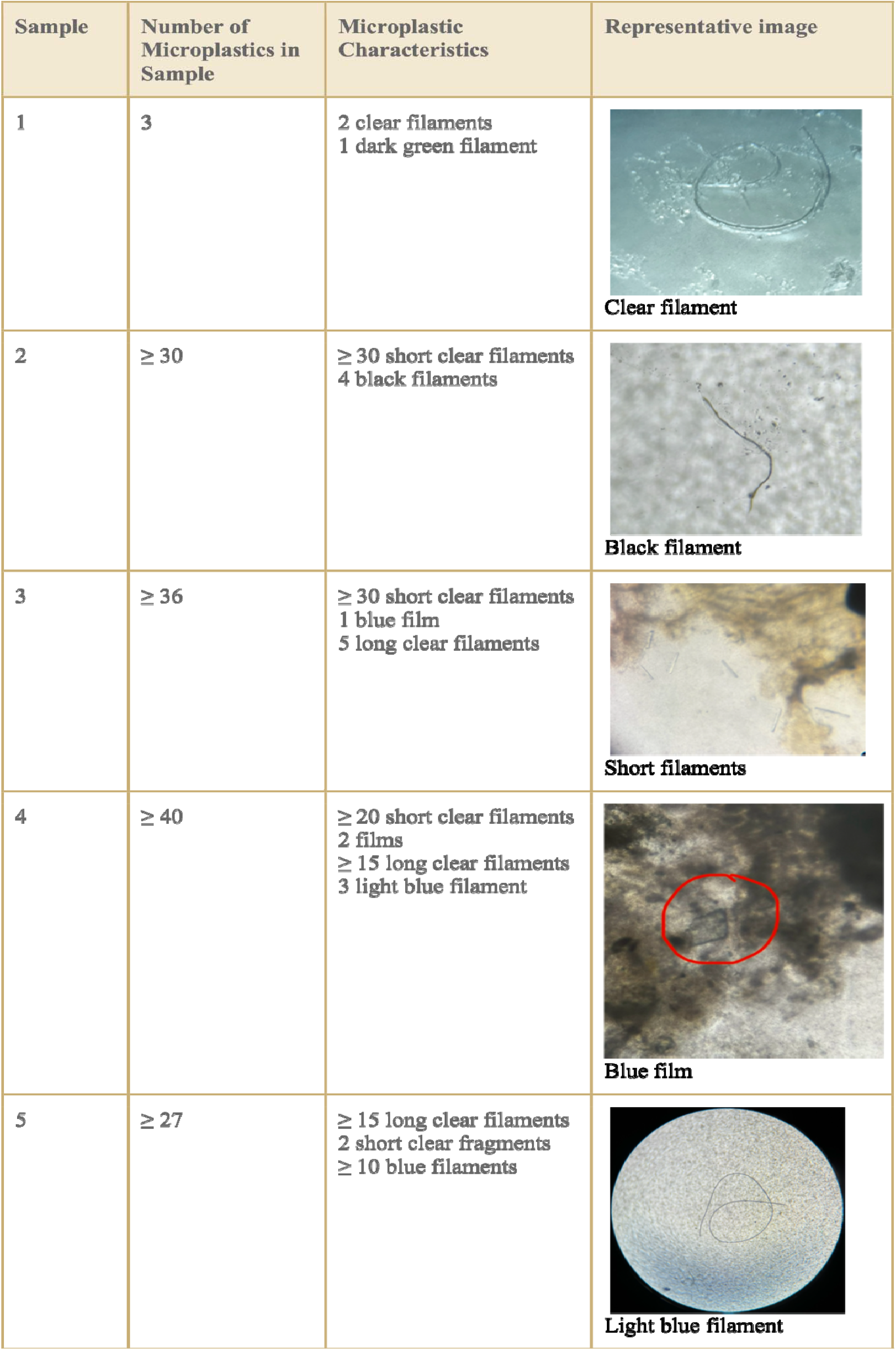
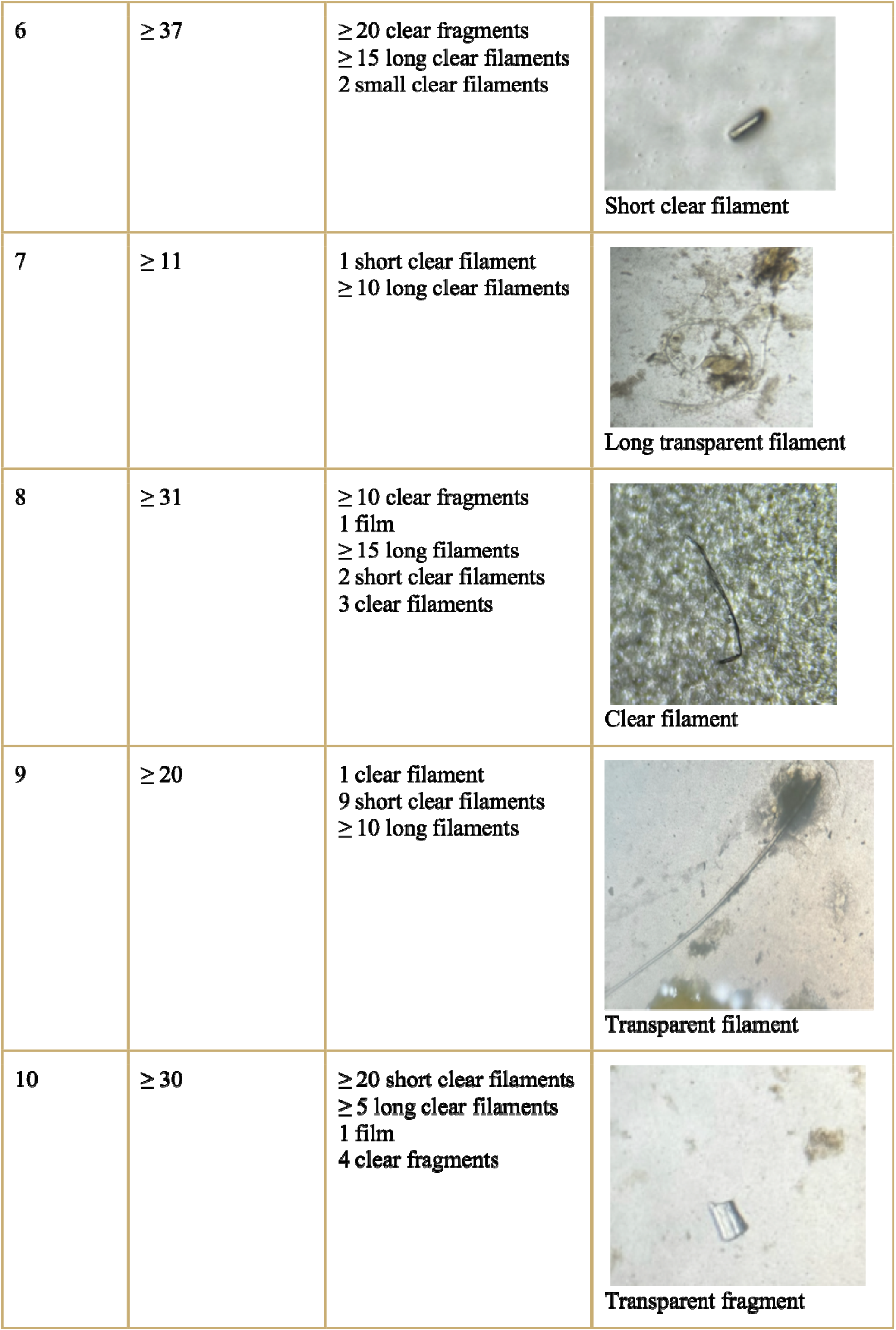
Anthropogenic Debris Recovered from Study Samples. The table above shows the sample number, total number of anthropogenic debris, the proportion of specific microplastic types, and an example image.

| Sample | Number of Microplastics in Sample | Microplastic Characteristics | Representative image |
| --- | --- | --- | --- |
| 1 | 3 | 2 clear filaments<br>1 dark green filament | Clear filament |
| 2 | $\geq 30$ | $\geq 30$ short clear filaments<br>4 black filaments | Black filament |
| 3 | $\geq 36$ | $\geq 30$ short clear filaments<br>1 blue film<br>5 long clear filaments | Short filaments |
| 4 | $\geq 40$ | $\geq 20$ short clear filaments<br>2 films<br>$\geq 15$ long clear filaments<br>3 light blue filament | Blue film |
| 5 | $\geq 27$ | $\geq 15$ long clear filaments<br>2 short clear fragments<br>$\geq 10$ blue filaments | Light blue filament |
| 6 | $\geq 37$ | $\geq 20$ clear fragments<br>$\geq 15$ long clear filaments<br>2 small clear filaments | Short clear filament |
| 7 | $\geq 11$ | 1 short clear filament<br>$\geq 10$ long clear filaments | Long transparent filament |
| 8 | $\geq 31$ | $\geq 10$ clear fragments<br>1 film<br>$\geq 15$ long filaments<br>2 short clear filaments<br>3 clear filaments | Clear filament |
| 9 | $\geq 20$ | 1 clear filament<br>9 short clear filaments<br>$\geq 10$ long filaments | Transparent filament |
| 10 | $\geq 30$ | $\geq 20$ short clear filaments<br>$\geq 5$ long clear filaments<br>1 film<br>4 clear fragments | Transparent fragment |

## Data Availability

All data produced in the present work are contained in the manuscript.

## Supplement

## MATERIALS

1. <u>12 petri dishes</u> (rinsed three times with ultrapure water before use to prevent contamination)
2. <u>12 glass jars</u> (rinsed three times with ultrapure water before use to prevent contamination)
3. <u>Glass stirrer</u> (rinsed three times with ultrapure water before use to prevent contamination)
4. <u>Beakers</u> (rinsed three times with ultrapure water before use to prevent contamination)
5. <u>20L ultrapure water</u> (for rinsing)
6. <u>Gloves</u> (to prevent contamination and harm)
7. <u>Goggles</u> (to prevent contamination and harm)
8. <u>Lab Coats</u> (to prevent contamination and harm)
9. <u>10% KOH solution</u> (to digest organic material)
10. <u>Water bath in Oven at 60°C</u> (to speed organic material decomposition)
11. <u>Laminar flow hood</u> (to prevent air contamination while sieving digested material)
12. <u>0.2mm sieve</u> (to sieve excess organic material to make looking at the sample easier)
13. <u>Spatula</u> (used to scrape leftover material into petri dishes after digestion)
14. <u>Dissection microscope</u> (looking at samples)
15. <u>TRAPDOOR Phone Adapter</u> (to take photos of the plastics found)
16. <u>Tape</u> (to mark samples)
17. <u>Sharpie</u> (to mark samples)

**Figure S1.**
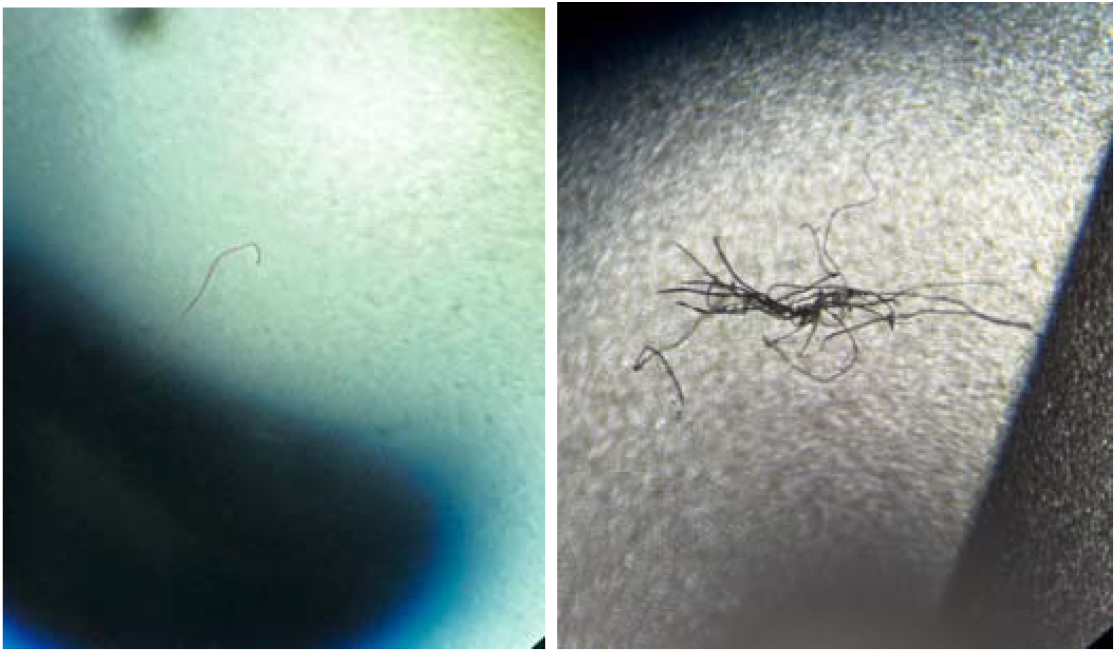
Anthropogenic debris noted in negative controls #1 and #2. Single episodes of anthropogenic debris found in controls #1 and #2. The picture on the left shows a red filament found in the ultrapure water control. The red filament was likely from a nearby red napkin. The image on the right displays a black filament from the laminar flow hood control. Control #3 was placed next to the microscope and was blank with no anthropogenic debris noted. Anthropogenic debris similar to control #2 was noted in sample 8 and was considered contaminant; no other samples contained similar debris.

